# Temporal pole initiation of neuronal avalanches is associated with episodic Memory performance in Mild Cognitive Impairment

**DOI:** 10.64898/2026.07.30.26359001

**Authors:** E. Gallo, M. Demuru, M. De Luca, E. Troisi Lopez, C. Granata, R. Nappo, M.C. Corsi, G. Sorrentino, D. Depannemaecker, M. Angiolelli, P. Sorrentino

## Abstract

Episodic-memory impairment is a defining feature of Mild cognitive impairment (MCI), yet the large-scale neural processes through which medial temporal pathology translates into poor cognitive performance remain unclear. Fast brain activity can be organized into transient, aperiodic bursts—neuronal avalanches—that propagate from hippocampal and adjacent temporal regions across distributed brain networks, potentially indexing interactions relevant to memory. We therefore hypothesized that episodic-memory impairment in MCI reflects an altered ability of the temporal pole to initiate these activity cascades. We analyzed resting-state, source-reconstructed MEG recordings from 29 individuals with MCI and 32 healthy controls (HC). Large-scale dynamics were described in terms of neuronal avalanches, and we quantified each temporal-pole region’s propensity to act as an “avalanche starter” —that is, to be the first region to become active. We then related this measure to episodic-memory performance and hippocampal volume. Although temporal-pole starter frequency did not differ between groups, its relationship with memory performance was reversed. Greater avalanche initiation from the left temporal pole was associated with poorer memory performance in MCI, as measured by the delayed free recall in the Free and Cued Selective Reminding Test. The same relationship was positive in HC. Within the MCI group, greater left-temporal-pole starter frequency was also associated with hippocampal atrophy. These findings suggest that MCI involves a qualitative reorganization of temporal-pole–initiated dynamics rather than a simple change in their frequency. This framework links local structural vulnerability to altered whole-brain dynamics and episodic-memory impairment.

**Significance Statement:** Memory decline is a hallmark of Mild Cognitive Impairment (MCI), often a precursor to Alzheimer’s disease, yet how the brain’s large-scale dynamics change to produce this decline remains unclear. Using magnetoencephalography, we studied brief bursts of coordinated activity — neuronal avalanches — and introduced the “avalanche starter”: a region’s propensity to initiate, rather than join, one of these cascades. We focused on the temporal pole, among the earliest regions affected by Alzheimer’s pathology. In healthy adults, frequent avalanche initiation there was linked to better memory; in MCI, the same pattern was linked to worse memory and hippocampal atrophy. This reversal shows that early cognitive decline reflects a qualitative reorganization of temporal-pole-driven dynamics, rather than a simple change in activity levels.

## 1. Introduction

Mild cognitive impairment (MCI) is commonly associated with an increased risk of dementia. Clinically, MCI is characterized by an objective decline in cognitive domains—most commonly episodic memory—while daily functioning is relatively preserved, with the amnestic subtype (aMCI) showing strong association with progression to Alzheimer’s disease (AD) (Celone et al., 2006; Petersen, 2004; Petersen et al., 2018; Raskin et al., 2015). Neuropathologically, AD pathology follows a hierarchical sequence: Braak’s staging scheme shows that amyloid and tau pathology first involve the medial temporal lobe—including the entorhinal cortex and temporal pole—before spreading to the cortex (Braak and Braak, 1995; Gold et al., 2001; Jacini et al., 2018a; Sorrentino et al., 2021b). Beyond structural atrophy (Du et al., 2001; Edmonds et al., 2020; Tabatabaei-Jafari et al., 2015), alterations in resting-state functional networks in temporal pole regions are established markers of disease progression (Apostolova et al., 2006; Burgess et al., 2002; Dickerson et al., 2005; Fornito et al., 2015; Friston, 2011; Jacini et al., 2018b; Lombardi et al., 2020; Sperling et al., 2010; Stoub et al., 2005; Wang et al., 2006; Yavuz et al., 2007). However, findings in MCI remain inconsistent across studies (Contreras et al., 2019; Jacini et al., 2018b; López et al., 2017; López-Sanz et al., 2017). Standard methods in functional connectivity rely on time-aggregated activities and assumptions of stationarity, which are at odds with a vast body of literature showing the complex, multimodal nature of brain dynamics (Zalesky et al., 2014). Furthermore, these traditional stationarity-based methods have been shown to replicate poorly. In contrast, large-scale neural activities are characterized by intermittent, aperiodic, scale-free bursts known as neuronal avalanches, typical of systems operating near criticality (Haldeman and Beggs, 2005; Shriki et al., 2013; Tagliazucchi et al., 2012). Critical dynamics support optimal cognitive flexibility (Cocchi et al., 2017), whereas deviations from criticality characterize neurological disorders (Cipriano et al., 2024; Corsi et al., 2024b; Polverino et al., 2022, 2024; Romano et al., 2023; Sorrentino et al., 2021a). In MCI, brain dynamics become less flexible, showing a reduced functional repertoire of neuronal avalanches in source-reconstructed MEG data (Sorrentino et al., 2021b). Furthermore, avalanche transition matrices (ATMs), which quantify propagation probabilities between regions, exhibit altered topology in MCI that predicts cognitive impairment beyond demographic variables and hippocampal volume (Sorrentino et al., 2021c). Despite this evidence, the local mechanisms altering cortico-hippocampal dynamics remain elusive. In temporal lobe epilepsy, structural alterations in the temporal pole have been linked to altered avalanche spreading and cognitive performance (Duma et al., 2023a, 2023b). By analogy, we hypothesized that the capacity of temporal pole regions to trigger activity cascades across networks—defined as avalanche “starter” dynamics—is altered in MCI. Specifically, given that avalanche propagation topology relates to cognitive performance (Corsi et al., 2024a; Duma et al., 2023a), we hypothesized that the propensity of the temporal pole to initiate avalanches would inversely relate to memory performance and decrease with structural atrophy. To test this hypothesis, we analyzed source-reconstructed MEG data in MCI patients and healthy controls (Romano et al., 2023), assessing temporal pole starter frequency and its associations with episodic memory and structural measures.

## 2. Material and Methods

### 2.1. Experimental Design

Thirty-one individuals with MCI (16 males and 15 females) were recruited from the Hermitage-Capodimonte clinic in Naples, Italy (age mean = 70.79, sd = 6.70; education level mean = 10.35, sd = 4.35). All participants were right-handed and native Italian speakers. MCI diagnosis was established according to the National Institute on Aging-Alzheimer’s Association criteria (Albert et al., 2011). Inclusion criteria were: (1) absence of neurological or systemic conditions potentially affecting cognitive status, (2) no contraindications to MRI or MEG recording, and (3) Fazekas score ≤2 (Fazekas et al., 1987) for both periventricular and deep white matter. Thirty-two healthy participants (19 males and 13 females), matched for age and years of education (age mean = 69.91, sd = 5.62; education level mean = 12.96, sd = 4.56), were enrolled as a control group (HC). All participants underwent neurological examination and MEG recording, as well as a comprehensive neuropsychological assessment covering the following cognitive domains: executive functions, sustained attention, verbal and visual memory, language abilities, praxic capacity, and reasoning. The study protocol was approved by the ‘‘Comitato Etico Campania Centro’’ (Prot.n.93C.E./Reg. n.14-17OSS) and all participants provided written informed consent in accordance with the Declaration of Helsinki. Cognitive functioning and comprehension abilities in MCI patients were sufficient to understand the aims of the study, allowing them to sign the consent form autonomously, without the need for a legal guardian or representative

### 2.2. MEG acquisition and preprocessing

All data were acquired using a magnetoencephalography (MEG) system. The MEG is equipped with 154 superconducting magnetometers and 9 reference sensors (Bonavolontà et al., 2025). The acquisition took place in a magnetically shielded room (ATB, Biomag, ULM, Germany) to reduce external noise. To locate the head position under the helmet, we used Fastrack (Polhemus), which digitized the positions of 4 anatomical landmarks (nasion, right and left preauricular points, and the vertex of the head) and 4 reference coils. Subjects were recorded twice (3,5 minutes each), with a one-minute break in the resting state. Subjects were asked to have their eyes closed. During the acquisition, we also recorded cardiac activity and eye movements to remove any physiological artifacts. MEG data preprocessing was performed similarly to Romano et al. (2023). Briefly, raw signals were filtered using a fourth-order Butterworth IIR band-pass filter implemented in the FieldTrip toolbox (Oostenveld et al., 2011) in MATLAB. Environmental noise was subsequently reduced using principal component analysis (PCA). Finally, a supervised independent component analysis (ICA) was applied to remove physiological artifacts: one component per participant was removed based on electrocardiography (ECG), while electrooculography (EOG)-related components were identified (rarely) and removed.

### 2.3. Source reconstruction

To reconstruct the time series of the regions of interest (ROIs), we relied on the automatic anatomical labeling atlas (AAL; Gong et al., 2009). The right temporal pole was defined as a composite region comprising the Right Middle Temporal Pole, Right Superior Temporal Pole, Right Parahippocampal Gyrus, and Right Hippocampus, while the left temporal pole was defined as a composite region comprising the Left Middle Temporal Pole, Left Superior Temporal Pole, Left Parahippocampal Gyrus, and Left Hippocampus. We conducted analyses both on each of these individual ROIs and on the composite right and left temporal pole, the latter defined as the mean across their constituent ROIs. These composite measures are hereafter referred to as the Right Temporal Pole Mean and Left Temporal Pole Mean, respectively.

Source reconstruction was performed by applying a linearly constrained minimum variance (LCMV) beamformer algorithm (Van Veen et al., 1997), accounting for volume conduction effects as described in Nolte (Nolte, 2003), and using each participant’s native MRI as the individual anatomical reference. Prior to source reconstruction, a trained operator manually inspected each participant’s structural MRI to identify the four anatomical fiducials recorded on the scalp before MEG acquisition — namely, the nasion, left preauricular, right preauricular, and vertex points. These landmarks were used to co-register the MEG data with the corresponding structural brain image, and source parcellation was carried out according to the AAL atlas definitions. The resulting ROI time series were subsequently decomposed into five canonical frequency bands: delta (0.5–4 Hz), theta (4–8 Hz), alpha (8–13 Hz), beta (13–30 Hz), and gamma (30–48 Hz).

### 2.5. Analysis of brain dynamics

Magnetoencephalography (MEG) data were acquired from each participant and organized in accordance with the Brain Imaging Data Structure (BIDS) convention (Gorgolewski et al., 2016). For each subject, all available runs were loaded and concatenated along the time axis. Since recording lengths varied across subjects, we excluded 3 subjects whose recording duration fell substantially below the group range, retaining only those whose duration was within an acceptable range of the majority, to ensure uniform dimensions across the group prior to further analysis.

To identify the spatial and temporal patterns of brain activity, MEG source signals were first normalized by converting each regional time series into z-scores, yielding signals with zero mean and standard deviation (SD). Under the assumption that brain activity arises from the superposition of many independent processes, signal amplitudes would be expected to follow a Gaussian distribution. However, MEG recordings systematically deviate from Gaussianity because of spatiotemporal correlations and collective neuronal activity (Fig. 1a). Comparison between the empirical amplitude distribution and its best-fitting Gaussian showed that the two distributions begin to diverge at approximately ±2.5 standard deviations. Based on this observation and previous studies (Shriki et al., 2013), a threshold of *θ*=±2.5 was adopted to identify significant neural events (Fig. 1b). We detected all continuous periods during which at least one ROI exhibited activity above the selected threshold. More specifically, an event started when the first suprathreshold activation appeared in any ROI and ended when the activity of all ROIs returned below threshold, such that each event was always preceded and followed by a silent interval. The extracted events displayed the characteristic statistical signatures of neuronal avalanches. In particular, the distributions of avalanche sizes and durations were compatible with an approximate power-law scaling, as expected for neuronal activity operating close to a critical regime. Avalanche size was defined as the cumulative sum of the suprathreshold activity across all active ROIs over the entire duration of the avalanche. These distributions are reported in the Supplementary Material (Figure S1).

**Figure 1.**
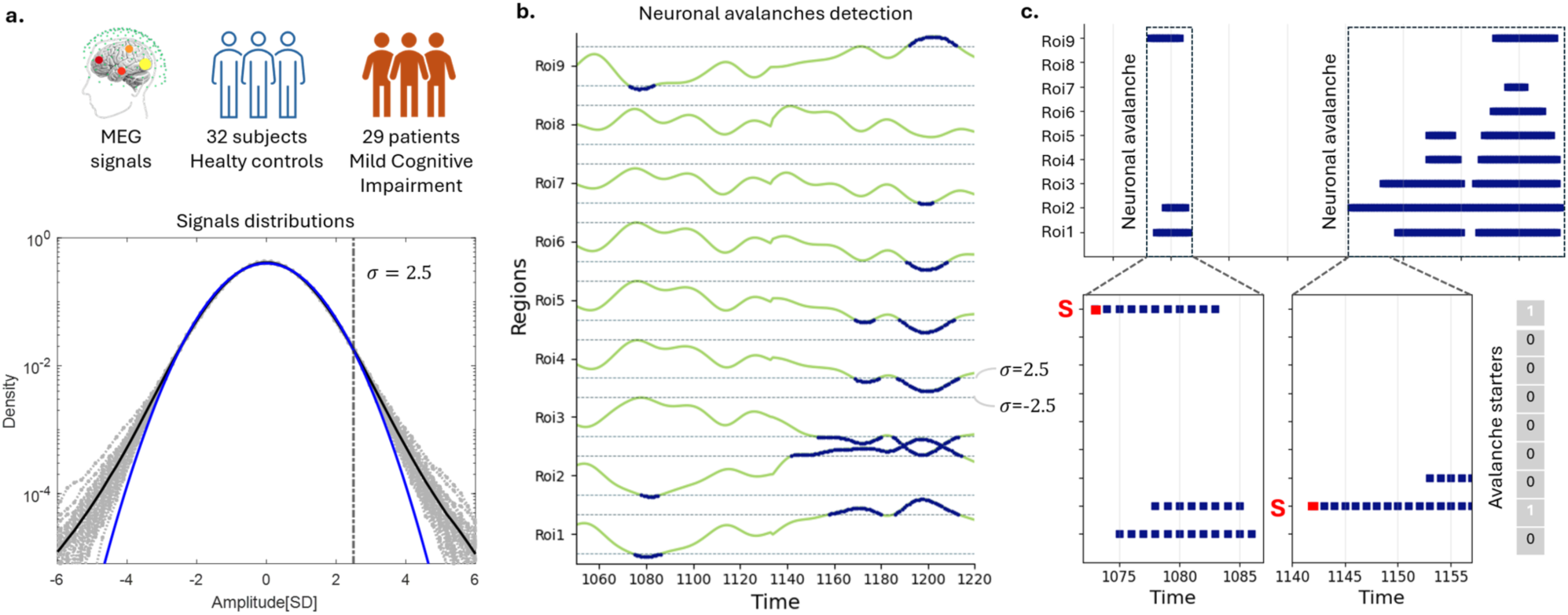
Pipeline overview. **(a)** Resting-state MEG recordings from 32 healthy controls (HC) and 29 patients with mild cognitive impairment (MCI) were source reconstructed into regional time series. For each subject, the time series were independently *z*-scored across time. The lower panel shows, as an example, the amplitude distribution obtained from the concatenated signals for each of the 32 healthy controls (gray lines). The blue curve represents the theoretical Gaussian distribution expected after *z*-scoring, whereas the black curve corresponds to the empirical mean distribution across subjects. The deviation of the empirical distribution from the Gaussian tails identifies the threshold of ∣σ∣>2.5 used to define suprathreshold events. **(b)** Avalanche detection across ROIs over time. Neuronal avalanches were defined as continuous periods during which at least one brain region exhibited suprathreshold activity. An avalanche starts with the first suprathreshold event and terminates when activity in all regions returns below threshold. Blue indicates the time points at which activities were suprathreshold. **(c)** Binary representation of a neuronal avalanche and identification of the starter region. For each avalanche, the starter region (red **S**) was identified as the region exhibiting the first suprathreshold activation (e.g., Roi 9 in the first avalanche shown and Roi 2 in the second avalanche shown). This procedure ultimately yields a vector with length equal to the number of ROIs, in which each entry corresponds to how many times an ROI acted as an “avalanche starter”. Then, the probability of each region acting as an avalanche starter was computed as the number of avalanches initiated by that region divided by the total number of avalanches in which the region was recruited at any point during the avalanche.

For each avalanche, we identified its starter, defined as the ROI in which the first suprathreshold activation occurred following the procedure described in (Angiolelli et al., 2025). We then counted the number of times each ROI acted as an avalanche starter throughout the recording and normalized this value by the total number of avalanches in which that ROI was recruited at any time during the propagation of the avalanche (Fig. 1c). This quantity, therefore, represents the probability that a region initiates an avalanche, conditional on its participation in the avalanche itself: P*_i_* = P(ROI *i* is starter∣ROI *i* participates).

### 2.6. MRI acquisition

Structural MRI data from both MCI patients and HC were acquired with a 3T Biograph mMR tomograph (Siemens Healthcare, Erlangen, Germany), using a 12-channel head coil. The acquisition protocol included three sequences: (i) a three-dimensional T1-weighted magnetization-prepared rapid acquisition gradient-echo sequence (240 sagittal planes, 214 × 21 mm² field of view, voxel size 1 × 1 × 1 mm³, TR/TE/TI 2400/2.5/1000 ms, flip angle 8°); (ii) a three-dimensional T2-weighted sampling perfection with application-optimized contrasts using different flip angle evolution sequence (SPACE, 240 sagittal planes, 214 × 214 mm² field of view, voxel size 1 × 1 × 1 mm³, TR/TE 3370/563 ms); and (iii) a two-dimensional T2-weighted turbo spin echo fluid-attenuated inversion recovery sequence (44 axial planes, 230 × 230 mm² field of view, voxel size 0.9 × 0.9 × 0.9 mm³, TR/TE/TI 9000/95/25,00, flip angle 150°). Volumetric analysis was carried out using FreeSurfer software (version 6.0) (FreeSurfer, 2012). Hippocampal volumes were normalized to the estimated total intracranial volume, while vascular burden was assessed using the Fazekas scale (Fazekas et al., 1987). Two patients and ten HCs either refused or did not complete the MRI scan; for these subjects, a standard MRI template was used for MEG source reconstruction, and they were consequently excluded from the structural volumetric analyses.

### 2.7. General Cognitive Assessment

Patients and healthy controls underwent a comprehensive neuropsychological assessment battery. General cognitive functioning was evaluated using the Mini Mental State Examination (Folstein et al., 1975), while executive functions were assessed by means of the Frontal Assessment Battery (Aiello et al., 2022)FAB; (Aiello et al., 2022).The Mental Deterioration Battery (MDB; Carlesimo et al., 1996) was administered to assess multiple cognitive domains, including episodic verbal memory (Rey Auditory Verbal Learning Test, RAVLT), verbal fluency (Phonemic Verbal Fluency test), language production (Sentence Construction test), non-verbal reasoning (Raven’s Coloured Progressive Matrices, 47 items), immediate visual memory (Immediate Visual Memory test), and visuoconstructional abilities (Constructional Praxis test, including freehand copying of drawings and copying with landmarks). Given that episodic memory impairment is a hallmark feature of mild cognitive impairment, participants additionally completed the Free and Cued Selective Reminding Test (Grober et al., 2010; Frasson et al., 2011) FCSRT; (Grober et al., 2010; Frasson et al., 2011) to specifically assess this domain. Finally, depressive symptomatology was screened using the Beck Depression Inventory (Beck et al., 1961) in all participants. Demographic and neuropsychological data are summarized in Table 1.

**Table 1:** Neuropsychological evaluation.

| **Test** | **MCI (n = 29)**<br>*Mean (± SD)* | **HC (n = 32)**<br>*Mean (± SD)* | **p value** |
| --- | --- | --- | --- |
| **BDI** | 9.66 (±6.66) | 7.81 (±4.44) | NS |
| **MMSE** | 26.43 (±1.68) | 27.57 (±1.7) | 0.0112 |
| **FAB** | 15.99 (±2.16) | 16.22 (±1.28) | NS |
| **RAY 15-WORD IMMEDIATE RECALL** | 26.97 (±5.88) | 44.86 (±6.05) | <0.0001 |
| **RAY 15-WORD DELAYED RECALL** | 4.19 (±1.59) | 10.23 (±2.46) | <0.0001 |
| **WORD FLUENCY** | 29.23 (±9.77) | 38.04 (±10.72) | 0.0017 |
| **PHRASE CONSTRUCTION** | 17.18 (±6.37) | 18.19 (±6.07) | NS |
| **47 RAVEN MATRICES** | 24.31 (±5.6) | 28.49 (±4.0) | 0.0019 |
| **IMMEDIATE VISUAL MEMORY** | 18.1 (±3.29) | 19.71 (±1.74) | 0.0256 |
| **FREEHAND COPYING OF DRAWINGS** | 9.21 (±2.14) | 10.04 (±1.24) | NS |
| **CONSTRUCTIVE APRAXIA WITH LANDMARKS** | 66.37 (±4.05) | 67.89 (±3.67) | NS |
| **FCRST** |  |  |  |
| **IMMEDIATE FREE RECALL** | 22.99 (±7.46) | 30.36 (±2.78) | <0.0001 |
| **DELAYED FREE RECALL** | 5.24 (±3.61) | 10.33 (±1.21) | <0.0001 |
| **IMMEDIATE TOTAL RECALL** | 32.45 (±4.66) | 35.94 (±0.25) | 0.0004 |
| **DELAYED TOTAL RECALL** | 9.14 (±3.49) | 11.94 (±0.25) | 0.0002 |
| **INDEX OF SENSITIVITY OF CUEING** | 0.78 (±0.23) | 0.99 (±0.03) | <0.0001 |

### 2.8. Statistical Analysis

Statistical Analysis was performed through Python 3.14.3, using the SciPy Python package. Based on the a priori hypothesis that temporal pole activity relates to episodic memory encoding and retrieval processes, Spearman’s rank correlations were computed between starter frequency per temporal pole ROIs and the five subscores of the Free and Cued Selective Reminding Test (FCSRT): Immediate Free Recall, Delayed Free Recall, Immediate Total Recall, Delayed Total Recall, and Index of Sensitivity to Cueing. The statistical significance threshold was set at *P* < 0.05. All p-values were corrected for multiple comparisons using the False Discovery Rate (Benjamini and Hochberg, 1995); unless otherwise specified, all p-values reported in the text refer to FDR-corrected values.

### 2.9. Code Accessibility

The code used for this analysis is publicly available in our GitHub repository at https://github.com/enricagallo001-ai/temporal_pole_MCI.git.

## 3. Results

Since our significant findings were restricted to the left hemisphere, from this point onward all regional references refer to left-hemisphere regions unless otherwise specified.

### 3.1. Cohort characteristics

MCI patients and HC showed no significant differences in age (HC: mean=69.91, sd=5.62; MCI: mean=70.79, sd=6.70) or education level (HC: mean=12.96, sd=4.56; MCI: mean=10.35, sd=4.35). From the initial MCI cohort of 32 patients, 4 were excluded due to acquisition lengths that were too short relative to the rest of the recordings, resulting in a final sample of 29 MCI patients and 32 HC. Neuropsychological evaluation revealed highly significant differences in memory performance between MCI patients and healthy controls, with MCI subjects consistently showing markedly lower scores across most measures, including both the FRCST and the RAVLT (Fig. 2).

**Figure 2.**
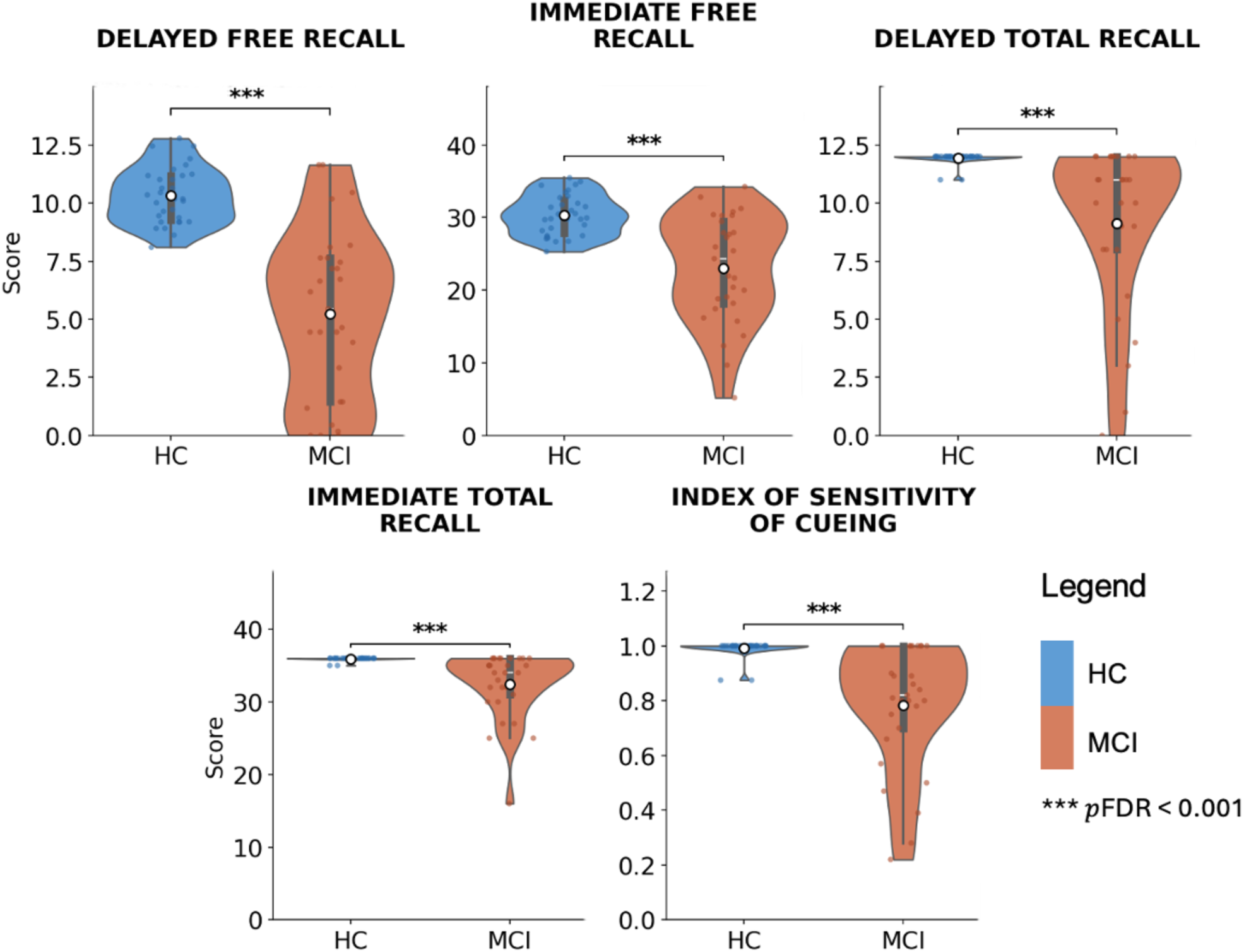
Memory Test Performance Comparison Between MCI Patients and Healthy Controls. Violin plots illustrating the distribution of neuropsychological memory test scores in healthy controls (HC, blue) and patients with Mild Cognitive Impairment (MCI, orange) across seven cognitive measures. Each violin displays the full score distribution, with the embedded box plot indicating the Mean (white dot), interquartile range, and 95% confidence interval. Individual subject scores are overlaid as jittered data points. MCI patients exhibited markedly lower performance compared to HC across all assessed measures.

### 3.2. Differences in starter frequencies

None of the ROIs examined showed statistically significant differences between MCI and HC after correction for multiple comparisons. Specifically, no significant differences emerged for Right Middle Temporal Pole (p_FDR = 0.865), Left Middle Temporal Pole (p_FDR = 0.865), Left ParaHippocampal Gyrus (p_FDR = 0.880), Right Hippocampus (p_FDR = 0.880), Left Superior Temporal Pole (p_FDR = 0.880), Right Superior Temporal Pole (p_FDR = 0.880), Right ParaHippocampal Gyrus (p_FDR = 0.880), or Left Hippocampus (p_FDR = 1.000). Similarly, at the lobe level, no significant differences were found for the right (p_FDR = 0.551) or left (p_FDR = 0.635) temporal pole mean.

### 3.3. Correlation between Temporal Pole Avalanche starters and clinical parameters

As previously described, all subjects underwent a standardized neuropsychological battery to compare cognitive performance between HC and MCI groups. Notably, a significant negative correlation was found between left temporal pole starter frequency and performance at the Delayed Free Recall phase in the FCRST (r = -0.413, p = 0.0261). On specific ROI analysis, we found a negative correlation between the ROI Left Middle Temporal Pole and Performance on the Delayed Free Recall of the FCRST. (Fig. 3). We found a positive correlation in the HC between Delayed Free Recall phase in FCRST and starter frequency both for Left Temporal Pole Mean (r = 0.414, p =0.0261) and for Left Middle Temporal Pole (r = 0.369, p =0.0409).

**Figure 3.**
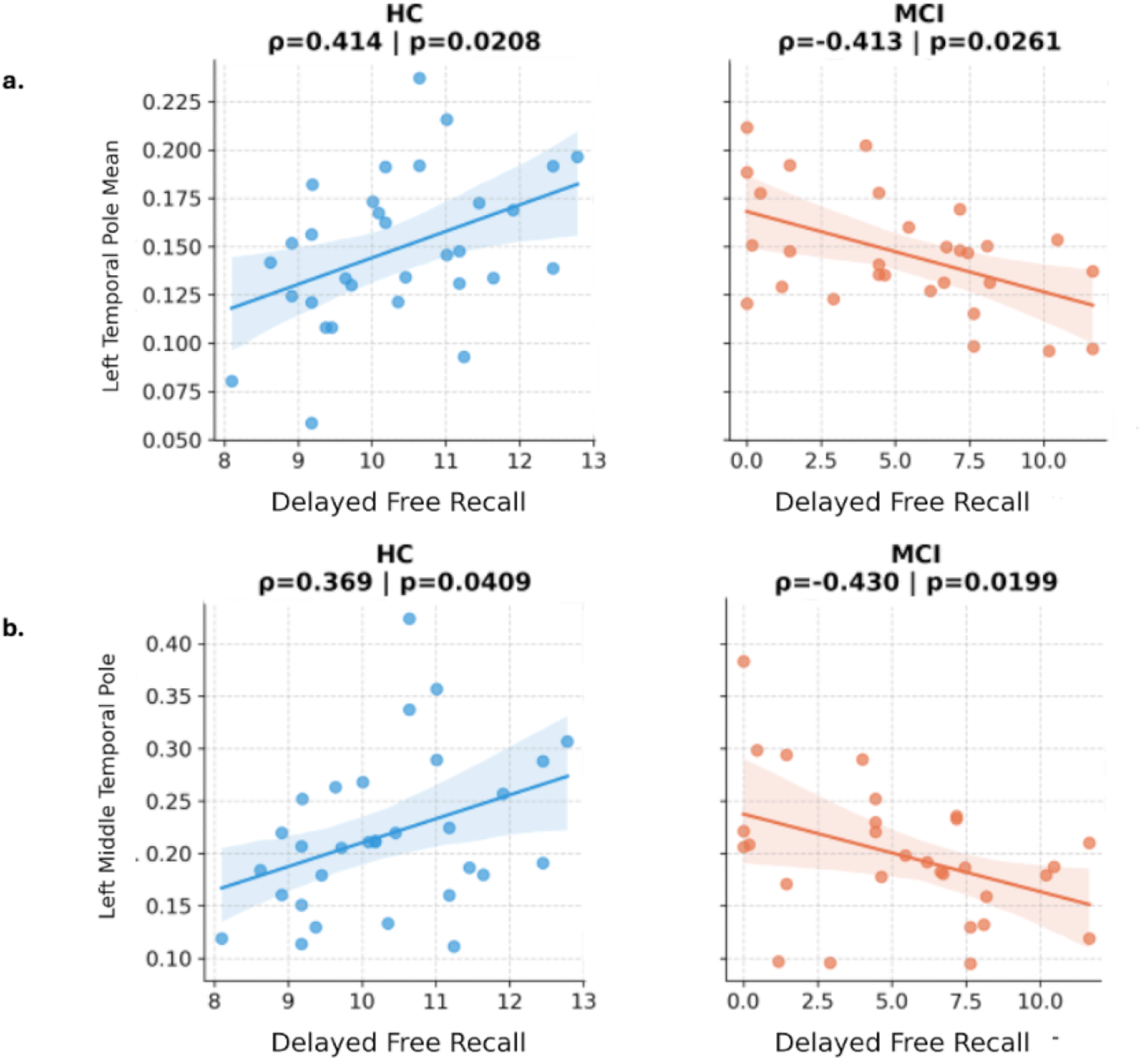
Spearman Correlations between FCRST - Delayed Free Recall and Avalanche Starters in Temporal Pole Areas. Scatter plots showing the correlation between avalanche starters and performance on the delayed recall phase of the Free and Cued Selective Reminding Test (FCSRT) in MCI patients and HC subjects, for the left temporal pole **(a)** and the left middle temporal pole **(b)**. In MCI patients, avalanche starters show an inverse correlation with delayed recall performance in the left temporal pole, and the same pattern holds for its subregion, the middle temporal pole. In HC subjects, this relationship is instead positive.

### 3.4. Correlation between Hippocampal Volumes and clinical parameters

A Spearman correlation analysis between FCSRT Delayed Recall and Left Hippocampal volume revealed a significant positive correlation (r = 0.471, p = 0.0132), indicating that lower hippocampal volumes were associated with poorer Delayed Recall performance in MCI patients (Fig. 4).

**Figure 4.**
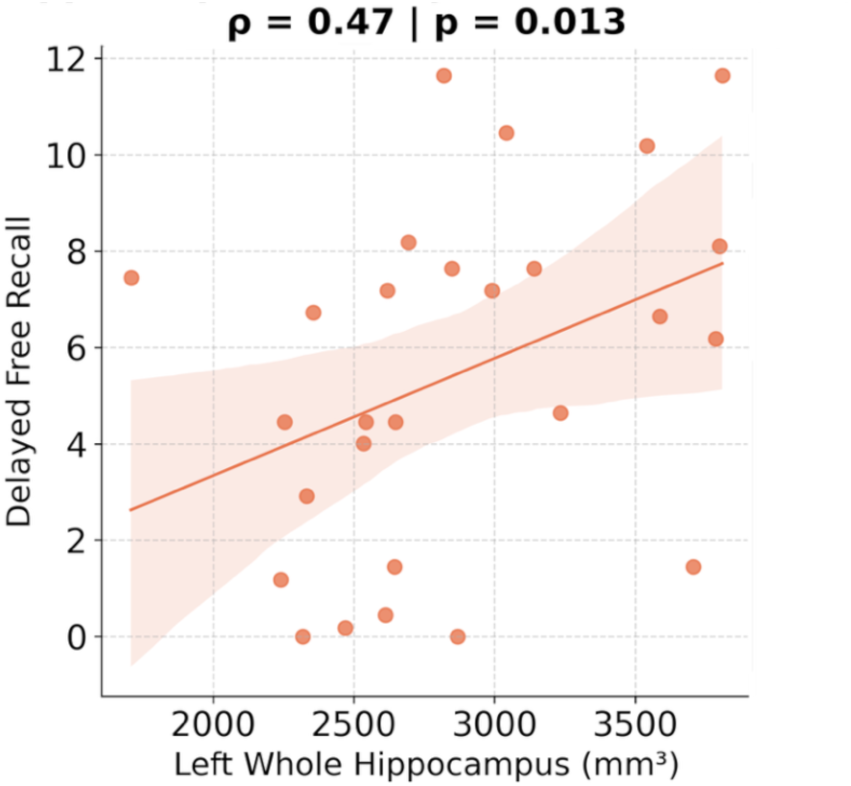
Spearman correlation between Left Hippocampal Volume and Delayed Free Recall scores in MCI. The scatter plot shows a significant positive correlation between left hippocampal volume and delayed recall performance on the FCSRT in MCI patients.

### 3.5. Correlation between Temporal Pole Avalanche Starters and Hippocampal volumes

We performed an additional correlation analysis between left hippocampal volume and Starter frequency in the Left Temporal Pole. This analysis was motivated by the well-documented occurrence of hippocampal atrophy in MCI and its established association with memory impairment. Results showed that, in the MCI population, left hippocampal volume was inversely correlated with left temporal pole Starter frequency (r = 0.0407, p = 0.0353) (Fig. 5).

**Figure 5.**
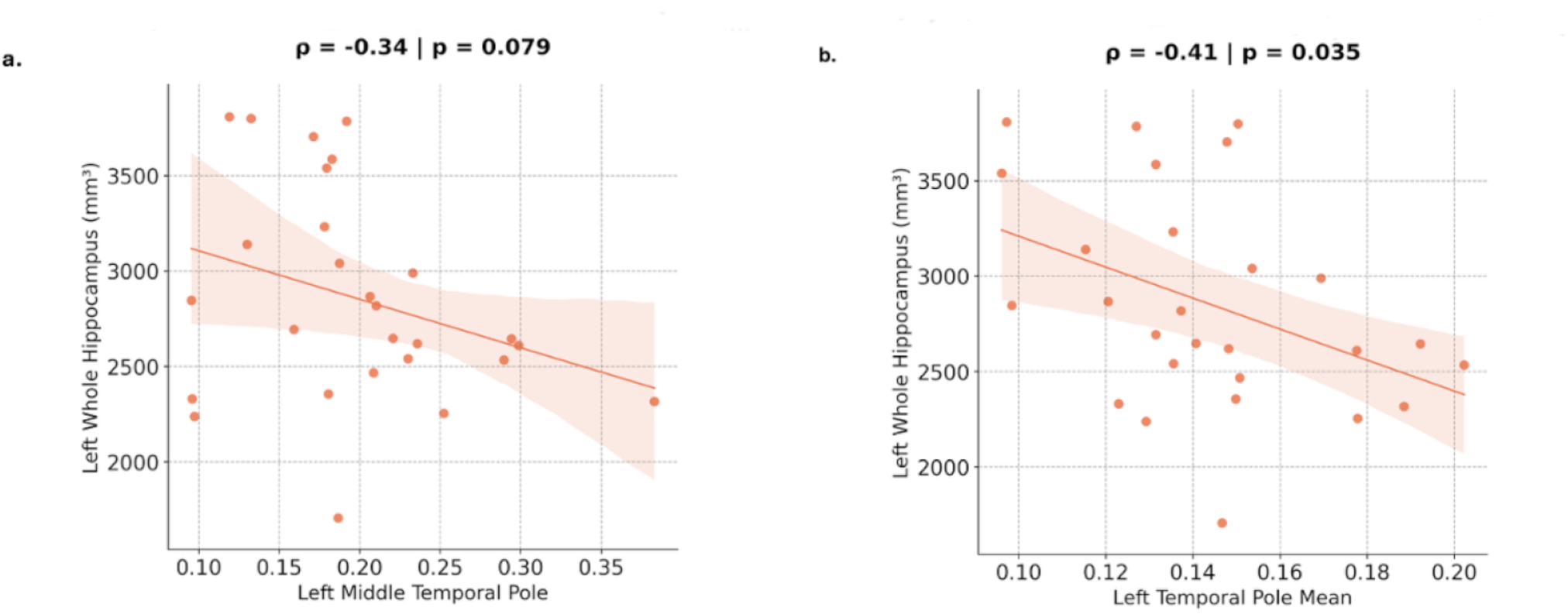
Spearman Correlation between Left Hippocampal Volumes and Left Temporal Pole Frequency Starters in MCI patients. Scatter plots showing the inverse correlations between avalanche starter frequency in the left temporal pole (a) or middle temporal pole (b) and left hippocampal volumes in MCI patients.

## 4. Discussion

In this manuscript, we set out to test whether the frequency of neuronal avalanches starting in the temporal pole changes in MCI and whether these changes index mnesic abilities and relate to degeneration. We found that, in MCI patients, the more the temporal poles start avalanches, the worse the mnesic performance. Increased starter frequency in the temporal poles is also related to brain atrophy (i.e., volume loss) in the hippocampus. The ability of the temporal poles to ignite avalanches did not differ between MCI individuals and controls. However, in the control group, we observed a significant direct relationship between the frequency at which the temporal poles act as avalanche starters and the mnesic performance. However, the different direction of the relationship observed in subjects with MCI, along with the correlation with brain atrophy, suggests that being an avalanche starter does not hold the same functional significance in MCI patients as in controls. In fact, the altered spreading of avalanches has been found to be modulated by a cognitive task (Corsi et al., 2024a) and by disease and altered brain structure (Duma et al., 2023b). Similarly, our results in healthy individuals suggest that avalanches originating in the temporal poles and then spreading across the brain might index large-scale interactions relevant to the performance of mnesic tasks. However, the opposite correlation between starter frequency in the temporal pole and mnesic performance observed in MCI might be interpreted as the manifestation of ineffective communication among areas. In fact, the more the hippocampus degenerates (lower volume), the more the temporal poles act as avalanche starters, and the worse the cognitive performance. In other words, since hippocampal volume is positively correlated with the FCRST, this pattern suggests that hippocampal volume loss may be accompanied by a compensatory increase in avalanche activity originating from the left temporal pole, as if to “offset” the loss of hippocampal contribution. However, in the healthy subjects, that is, in the absence of hippocampal damage, the temporal pole activity supports memory performance. We suggest that this disruption in the avalanche-starter dynamics may represent a neurophysiological substrate underlying the memory alterations observed clinically in MCI, linking network-level dynamics, in terms of neuronal avalanches, to the disease’s characteristic mnemonic deficits.

Taken together, these findings suggest that the same neural mechanism that operates adaptively during healthy aging becomes maladaptive once hippocampal damage is in place—possibly reflecting a loss of functional specificity in recruitment, rather than a simple quantitative increase in compensatory effort. Dickerson et al. (Dickerson et al., 2005b) found that greater hippocampal/MTL activation during a memory task, rather than reduced activation, predicted worse outcomes in MCI: “decliners” showed higher hippocampal activation than stable subjects, while “converters” to AD showed lower MTL activation than decliners. This suggests MTL hyperactivation reflects a compensatory but maladaptive mechanism, present early in decline but lost as patients approach dementia.

The left-hemispheric lateralization of these effects might be related to the nature of the cognitive domain probed by the FCRST, which is a verbal memory test. Indeed, the left temporal pole has been consistently implicated in the retrieval of lexical labels for concrete entities, with anatomically distinct sectors supporting word retrieval for different conceptual categories of items (Damasio et al., 1996). Building on this, left temporal pole activity has been shown to be specifically engaged during the retrieval of labels for unique, specific entities, rather than being restricted to a particular semantic category — a category-general property confirmed across multiple studies (Grabowski et al., 2001). This category-independence is particularly relevant to the present findings, as the FCRST requires the recall of specific verbal labels for items drawn from multiple semantic categories (e.g., living and non-living). The apparent category-general nature of left temporal pole engagement in label retrieval is therefore consistent with, and may help explain, its involvement across the heterogeneous item set used in the FCRST, further supporting the plausibility of a left-lateralized, verbally-mediated substrate for the avalanche dynamics observed in this task. Beyond its role in verbal label retrieval, the broader medial temporal lobe also appears to support both semantic and episodic memory through its involvement in verbal processing (Menon et al., 2002). Future investigations into temporal pole-initiated avalanche patterns could prove particularly informative. In particular, identifying the brain regions most frequently recruited in avalanches initiated by the left temporal pole in MCI patients may clarify whether these regions aberrantly engage areas not canonically associated with memory processing, potentially revealing a dysfunctional connectivity signature. Overall, these results point to avalanche-starter frequency in the left temporal pole as a candidate biomarker for early memory decline and its progression towards dementia.

## Author contributions

S.P., S.G., and T.L.E. designed research; D.L.M., T.L.E. and E.G. performed research; T.L.E, D.M. and A.M. contribuited unpublished reagents/analytic tools; E.G., D.M., and A.M. analyzed data; E.G. wrote the paper; D.L.M., D.M., T.L.E., G.C., N.R., C.M.C., S.G., D.D., A.M. and S.P., edited the paper.

## Data Availability

The data that support the findings of this study (including preprocessed MEG metrics, structural MRI volumes, and neuropsychological cognitive scores) are available from the corresponding author upon reasonable request from qualified researchers, subject to institutional data-use agreements and ethical guidelines.

## Acknowledgments

This work was supported by the Italian Government, Ministry of Enterprises and Made in Italy (formerly Ministry of Economic Development), under the Accordi per l’Innovazione program, project AUDACE (“Approccio User-friendly integrato per Diagnosi, Assistenza e Cura Efficaci” — User-friendly Integrated Approach for Effective Diagnosis, Assistance and Care), CUP: B69J23006050007.

This work was also supported by the European Union – NextGenerationEU, under the Italian National Recovery and Resilience Plan (PNRR), Investment 3.1, Mission 4, Component 2, project IR0000011, EBRAINS-Italy.

AI-assisted tools were used to support code development and documentation for this project.

## Supplementary Material

**Figure S1.**
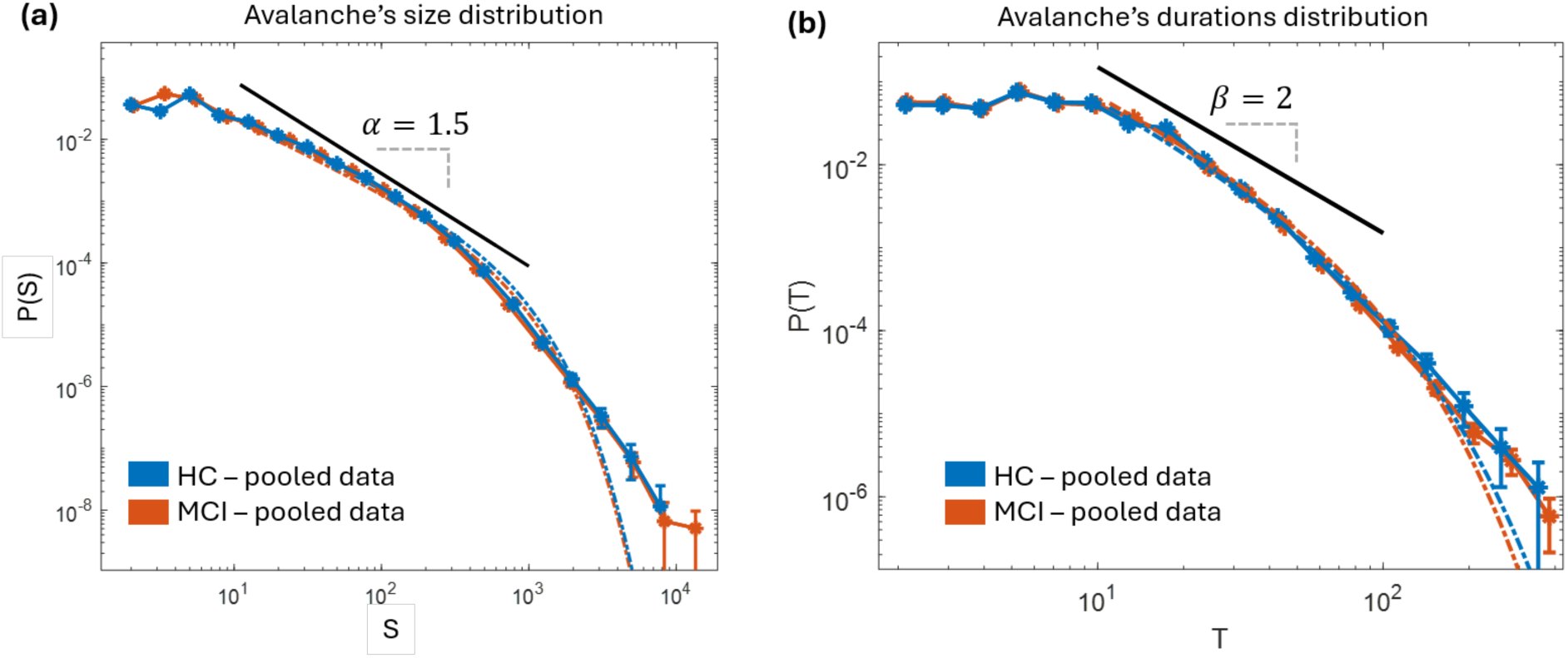
**(a)** Distribution of avalanche sizes pooled across all healthy controls (HC, blue) and patients with mild cognitive impairment (MCI, orange), displayed on logarithmic axes. Both distributions exhibit an approximately scale-free regime over an intermediate range of avalanche sizes (black line, slope corresponding to α=1.5 shown for visual guidance). The power-law exponent was estimated independently for the HC and MCI groups using (Alstott et al., 2014). When the fitting was restricted to the scaling range identified by the black line shown in the figure (S∈[10,1000]), the estimated exponents were α-HC≈1.43 and α-MCI≈1.45. For HC (MCI) group, the fit was characterized by a Kolmogorov–Smirnov distance of D=0.060 (D=0.064). The power-law model was strongly preferred over an exponential distribution (HC: log-likelihood ratio R=5000.53, *p*=2.4×10^(−25); MCI: R=9638.93, *p*=9.34×10^(−25)). When the upper cutoff *x_max_* was not taken into account, the truncated power-law was also significantly favored over a log-normal distribution (HC: R=220.46, *p*=4.44×10^(−5), MCI: *R*=810.12, *p*=4.65×10^(−15)). Error bars represent twice the estimated standard deviation of the empirical probability in each bin, assuming binomial counting statistics. Specifically, if *P*(*x*) denotes the empirical probability of observing an avalanche size within the interval [*x*,*x*+Δ*x*], the corresponding uncertainty was estimated as 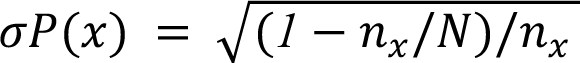, where *n_x_* is the number of avalanches falling in the interval [*x*,*x*+Δ*x*], and *N* is the total number of avalanches. Error bars correspond to (Scarpetta et al., 2023)*2σP*(*x*). (Scarpetta et al., 2023) **(b)** Distribution of avalanche durations pooled across all healthy controls (HC, blue) and patients with mild cognitive impairment (MCI, orange), displayed on logarithmic axes. As before, both distributions exhibit an approximately scale-free regime over an intermediate range of avalanche durations (black line, slope corresponding to β=2 shown for visual guidance). When the fitting was restricted to the scaling range identified by the black line shown in the figure (T∈[10,100]), the estimated exponents were β-HC≈2.36 and β-MCI≈2.38. For the HC (MCI) group, the fit was characterized by a Kolmogorov–Smirnov distance of D= 0.059 (D=0.053). The power-law model was strongly preferred over an exponential distribution (HC: R=197.820, *p*=0.001; MCI: R=172.23, *p*=0.001). When the upper cutoff *x_max_* was not taken into account, the truncated power-law was also significantly favored over a log-normal distribution (HC: R=55.35, *p*=7.93×10^(−23), MCI: R=291.72, *p*=7.6×10^(−12)).

## References

1. Aiello EN, Esposito A, Gramegna C, Gazzaniga V, Zago S, Difonzo T, Appollonio IM, Bolognini N (2022) The Frontal Assessment Battery (FAB) and its sub-scales: validation and updated normative data in an Italian population sample. Neurol Sci Off J Ital Neurol Soc Ital Soc Clin Neurophysiol 43:979–984.

2. Albert MS, DeKosky ST, Dickson D, Dubois B, Feldman HH, Fox NC, Gamst A, Holtzman DM, Jagust WJ, Petersen RC, Snyder PJ, Carrillo MC, Thies B, Phelps CH (2011) The diagnosis of mild cognitive impairment due to Alzheimer’s disease: recommendations from the National Institute on Aging-Alzheimer’s Association workgroups on diagnostic guidelines for Alzheimer’s disease. Alzheimers Dement J Alzheimers Assoc 7:270–279.

3. Alstott J, Bullmore E, Plenz D (2014) Powerlaw: a Python package for analysis of heavy-tailed distributions. PLoS ONE 9:e85777.

4. Apostolova LG, Dutton RA, Dinov ID, Hayashi KM, Toga AW, Cummings JL, Thompson PM (2006) Conversion of mild cognitive impairment to Alzheimer disease predicted by hippocampal atrophy maps. Arch Neurol 63:693–699.

5. Beck AT, Ward CH, Mendelson M, Mock J, Erbaugh J (1961) An inventory for measuring depression. Arch Gen Psychiatry 4:561–571.

6. Benjamini Y, Hochberg Y (1995) Controlling the False Discovery Rate: A Practical and Powerful Approach to Multiple Testing. J R Stat Soc Ser B Methodol 57:289–300.

7. Bonavolontà C, Vettoliere A, Sorrentino P, Granata C (2025) Superconducting Quantum Magnetometers for Brain Investigations. Sensors 25:4625.

8. Braak H, Braak E (1995) Staging of Alzheimer’s disease-related neurofibrillary changes. Neurobiol Aging 16:271–278; discussion 278-284.

9. Burgess N, Maguire EA, O’Keefe J (2002) The human hippocampus and spatial and episodic memory. Neuron 35:625–641.

10. Carlesimo GA, Caltagirone C, Gainotti G (1996) The Mental Deterioration Battery: normative data, diagnostic reliability and qualitative analyses of cognitive impairment. The Group for the Standardization of the Mental Deterioration Battery. Eur Neurol 36:378–384.

11. Celone KA, Calhoun VD, Dickerson BC, Atri A, Chua EF, Miller SL, DePeau K, Rentz DM, Selkoe DJ, Blacker D, Albert MS, Sperling RA (2006) Alterations in memory networks in mild cognitive impairment and Alzheimer’s disease: an independent component analysis. J Neurosci Off J Soc Neurosci 26:10222–10231.

12. Cipriano L, Minino R, Liparoti M, Polverino A, Romano A, Bonavita S, Pirozzi MA, Quarantelli M, Jirsa V, Sorrentino G, Sorrentino P, Troisi Lopez E (2024) Flexibility of brain dynamics is increased and predicts clinical impairment in relapsing-remitting but not in secondary progressive multiple sclerosis. Brain Commun 6:fcae112.

13. Cocchi L, Gollo LL, Zalesky A, Breakspear M (2017) Criticality in the brain: A synthesis of neurobiology, models and cognition. Prog Neurobiol 158:132–152.

14. Contreras JA, Avena-Koenigsberger A, Risacher SL, West JD, Tallman E, McDonald BC, Farlow MR, Apostolova LG, Goñi J, Dzemidzic M, Wu Y-C, Kessler D, Jeub L, Fortunato S, Saykin AJ, Sporns O (2019) Resting state network modularity along the prodromal late onset Alzheimer’s disease continuum. NeuroImage Clin 22:101687.

15. Corsi M-C, Sorrentino P, Schwartz D, George N, Gollo LL, Chevallier S, Hugueville L, Kahn AE, Dupont S, Bassett DS, Jirsa V, De Vico Fallani F (2024a) Measuring neuronal avalanches to inform brain-computer interfaces. iScience 27:108734.

16. Corsi M-C, Troisi Lopez E, Sorrentino P, Cuozzo S, Danieli A, Bonanni P, Duma GM (2024b) Neuronal avalanches in temporal lobe epilepsy as a noninvasive diagnostic tool investigating large scale brain dynamics. Sci Rep 14:14039.

17. Damasio H, Grabowski TJ, Tranel D, Hichwa RD, Damasio AR (1996) A neural basis for lexical retrieval. Nature 380:499–505.

18. Dickerson BC, Salat DH, Greve DN, Albert MS, Blacker D, Sperling RA (2005a) [P-095]: An fMRI investigation of memory-related medial temporal lobe activation in mild cognitive impairment prior to dementia. Alzheimers Dement 1:S38–S38.

19. Dickerson BC, Salat DH, Greve DN, Chua EF, Rand-Giovannetti E, Rentz DM, Bertram L, Mullin K, Tanzi RE, Blacker D, Albert MS, Sperling RA (2005b) Increased hippocampal activation in mild cognitive impairment compared to normal aging and AD. Neurology 65:404–411.

20. Du AT, Schuff N, Amend D, Laakso MP, Hsu YY, Jagust WJ, Yaffe K, Kramer JH, Reed B, Norman D, Chui HC, Weiner MW (2001) Magnetic resonance imaging of the entorhinal cortex and hippocampus in mild cognitive impairment and Alzheimer’s disease. J Neurol Neurosurg Psychiatry 71:441–447.

21. Duma GM, Danieli A, Mento G, Vitale V, Opipari RS, Jirsa V, Bonanni P, Sorrentino P (2023a) Altered spreading of neuronal avalanches in temporal lobe epilepsy relates to cognitive performance: A resting-state hdEEG study. Epilepsia 64:1278–1288.

22. Duma GM, Pellegrino G, Rabuffo G, Danieli A, Antoniazzi L, Vitale V, Scotto Opipari R, Bonanni P, Sorrentino P (2023b) Altered spread of waves of activities at large scale is influenced by cortical thickness organization in temporal lobe epilepsy: a magnetic resonance imaging–high-density electroencephalography study. Brain Commun 6:fcad348.

23. Edmonds EC, Weigand AJ, Hatton SN, Marshall AJ, Thomas KR, Ayala DA, Bondi MW, McDonald CR, Alzheimer’s Disease Neuroimaging Initiative (2020) Patterns of longitudinal cortical atrophy over 3 years in empirically derived MCI subtypes. Neurology 94:e2532–e2544.

24. Fazekas F, Chawluk JB, Alavi A, Hurtig HI, Zimmerman RA (1987) MR signal abnormalities at 1.5 T in Alzheimer’s dementia and normal aging. AJR Am J Roentgenol 149:351–356.

25. Folstein MF, Folstein SE, McHugh PR (1975) “Mini-mental state”. A practical method for grading the cognitive state of patients for the clinician. J Psychiatr Res 12:189–198.

26. Fornito A, Zalesky A, Breakspear M (2015) The connectomics of brain disorders. Nat Rev Neurosci 16:159–172.

27. Frasson P, Ghiretti R, Catricalà E, Pomati S, Marcone A, Parisi L, Rossini PM, Cappa SF, Mariani C, Vanacore N, Clerici F (2011) Free and Cued Selective Reminding Test: an Italian normative study. Neurol Sci Off J Ital Neurol Soc Ital Soc Clin Neurophysiol 32:1057–1062.

28. Friston KJ (2011) Functional and effective connectivity: a review. Brain Connect 1:13–36.

29. Gong G, He Y, Concha L, Lebel C, Gross DW, Evans AC, Beaulieu C (2009) Mapping Anatomical Connectivity Patterns of Human Cerebral Cortex Using In Vivo Diffusion Tensor Imaging Tractography. Cereb Cortex 19:524–536.

30. Gorgolewski KJ et al. (2016) The brain imaging data structure, a format for organizing and describing outputs of neuroimaging experiments. Sci Data 3:160044.

31. Grabowski TJ, Damasio H, Tranel D, Ponto LL, Hichwa RD, Damasio AR (2001) A role for left temporal pole in the retrieval of words for unique entities. Hum Brain Mapp 13:199–212.

32. Grober E, Sanders AE, Hall C, Lipton RB (2010) Free and cued selective reminding identifies very mild dementia in primary care. Alzheimer Dis Assoc Disord 24:284–290.

33. Haldeman C, Beggs JM (2005) Critical Branching Captures Activity in Living Neural Networks and Maximizes the Number of Metastable States. Phys Rev Lett 94:058101.

34. Jacini F, Sorrentino P, Lardone A, Rucco R, Baselice F, Cavaliere C, Aiello M, Orsini M, Iavarone A, Manzo V, Carotenuto A, Granata C, Hillebrand A, Sorrentino G (2018a) Amnestic Mild Cognitive Impairment Is Associated With Frequency-Specific Brain Network Alterations in Temporal Poles. Front Aging Neurosci 10 Available at: https://www.frontiersin.org/journals/aging-neuroscience/articles/10.3389/fnagi.2018.00400/full [Accessed July 7, 2026].

35. Jacini F, Sorrentino P, Lardone A, Rucco R, Baselice F, Cavaliere C, Aiello M, Orsini M, Iavarone A, Manzo V, Carotenuto A, Granata C, Hillebrand A, Sorrentino G (2018b) Amnestic Mild Cognitive Impairment Is Associated With Frequency-Specific Brain Network Alterations in Temporal Poles. Front Aging Neurosci 10 Available at: https://www.frontiersin.org/journals/aging-neuroscience/articles/10.3389/fnagi.2018.00400/full [Accessed April 22, 2026].

36. Jongsiriyanyong S, Limpawattana P (2018) Mild Cognitive Impairment in Clinical Practice: A Review Article. Am J Alzheimers Dis Other Demen 33:500–507.

37. Lombardi G, Crescioli G, Cavedo E, Lucenteforte E, Casazza G, Bellatorre A, Lista C, Costantino G, Frisoni G, Virgili G, Filippini G (2020) Structural magnetic resonance imaging for the early diagnosis of dementia due to Alzheimer’s disease in people with mild cognitive impairment. Cochrane Database Syst Rev 2020:CD009628.

38. López ME, Engels MMA, van Straaten ECW, Bajo R, Delgado ML, Scheltens P, Hillebrand A, Stam CJ, Maestú F (2017) MEG Beamformer-Based Reconstructions of Functional Networks in Mild Cognitive Impairment. Front Aging Neurosci 9 Available at: https://www.frontiersin.org/journals/aging-neuroscience/articles/10.3389/fnagi.2017.00107/full [Accessed July 22, 2026].

39. López-Sanz D, Bruña R, Garcés P, Martín-Buro MC, Walter S, Delgado ML, Montenegro M, López Higes R, Marcos A, Maestú F (2017) Functional Connectivity Disruption in Subjective Cognitive Decline and Mild Cognitive Impairment: A Common Pattern of Alterations. Front Aging Neurosci 9 Available at: https://www.frontiersin.org/journals/aging-neuroscience/articles/10.3389/fnagi.2017.00109/full [Accessed July 22, 2026].

40. Menon V, Boyett-Anderson JM, Schatzberg AF, Reiss AL (2002) Relating semantic and episodic memory systems. Cogn Brain Res 13:261–265.

41. Nolte G (2003) The magnetic lead field theorem in the quasi-static approximation and its use for magnetoencephalography forward calculation in realistic volume conductors. Phys Med Biol 48:3637–3652.

42. Oostenveld R, Fries P, Maris E, Schoffelen J-M (2011) FieldTrip: Open source software for advanced analysis of MEG, EEG, and invasive electrophysiological data. Comput Intell Neurosci 2011:156869.

43. Petersen RC (2004) Mild cognitive impairment as a diagnostic entity. J Intern Med 256:183–194.

44. Petersen RC, Lopez O, Armstrong MJ, Getchius TSD, Ganguli M, Gloss D, Gronseth GS, Marson D, Pringsheim T, Day GS, Sager M, Stevens J, Rae-Grant A (2018) Practice guideline update summary: Mild cognitive impairment. Neurology 90:126–135.

45. Polverino A, Troisi Lopez E, Liparoti M, Minino R, Romano A, Cipriano L, Trojsi F, Jirsa V, Sorrentino G, Sorrentino P (2024) Altered spreading of fast aperiodic brain waves relates to disease duration in Amyotrophic Lateral Sclerosis. Clin Neurophysiol 163:14–21.

46. Polverino A, Troisi Lopez E, Minino R, Liparoti M, Romano A, Trojsi F, Lucidi F, Gollo L, Jirsa V, Sorrentino G, Sorrentino P (2022) Flexibility of Fast Brain Dynamics and Disease Severity in Amyotrophic Lateral Sclerosis. Neurology 99:e2395–e2405.

47. Raskin J, Cummings J, Hardy J, Schuh K, Dean RA (2015) Neurobiology of Alzheimer’s Disease: Integrated Molecular, Physiological, Anatomical, Biomarker, and Cognitive Dimensions. Curr Alzheimer Res 12:712–722.

48. Romano A, Troisi Lopez E, Cipriano L, Liparoti M, Minino R, Polverino A, Cavaliere C, Aiello M, Granata C, Sorrentino G, Sorrentino P (2023) Topological changes of fast large-scale brain dynamics in mild cognitive impairment predict early memory impairment: a resting-state, source reconstructed, magnetoencephalography study. Neurobiol Aging 132:36–46.

49. Scarpetta S, Morisi N, Mutti C, Azzi N, Trippi I, Ciliento R, Apicella I, Messuti G, Angiolelli M, Lombardi F, Parrino L, Vaudano AE (2023) Criticality of neuronal avalanches in human sleep and their relationship with sleep macro- and micro-architecture. iScience 26:107840.

50. Shriki O, Alstott J, Carver F, Holroyd T, Henson RNA, Smith ML, Coppola R, Bullmore E, Plenz D (2013) Neuronal Avalanches in the Resting MEG of the Human Brain. J Neurosci 33:7079–7090.

51. Sorrentino P, Rucco R, Baselice F, De Micco R, Tessitore A, Hillebrand A, Mandolesi L, Breakspear M, Gollo LL, Sorrentino G (2021a) Flexible brain dynamics underpins complex behaviours as observed in Parkinson’s disease. Sci Rep 11:4051.

52. Sorrentino P, Rucco R, Lardone A, Liparoti M, Troisi Lopez E, Cavaliere C, Soricelli A, Jirsa V, Sorrentino G, Amico E (2021b) Clinical connectome fingerprints of cognitive decline. NeuroImage 238:118253.

53. Sorrentino P, Seguin C, Rucco R, Liparoti M, Troisi Lopez E, Bonavita S, Quarantelli M, Sorrentino G, Jirsa V, Zalesky A (2021c) The structural connectome constrains fast brain dynamics Behrens TE, Vidaurre D, Quinn AJ, O’Neill G, eds. eLife 10:e67400.

54. Sperling RA, Dickerson BC, Pihlajamaki M, Vannini P, LaViolette PS, Vitolo OV, Hedden T, Becker JA, Rentz DM, Selkoe DJ, Johnson KA (2010) Functional alterations in memory networks in early Alzheimer’s disease. Neuromolecular Med 12:27–43.

55. Stoub TR, Bulgakova M, Leurgans S, Bennett DA, Fleischman D, Turner DA, deToledo-Morrell L (2005) MRI predictors of risk of incident Alzheimer disease: a longitudinal study. Neurology 64:1520–1524.

56. Tabatabaei-Jafari H, Shaw ME, Cherbuin N (2015) Cerebral atrophy in mild cognitive impairment: A systematic review with meta-analysis. Alzheimers Dement 1:487–504.

57. Tagliazucchi E, Balenzuela P, Fraiman D, Chialvo DR (2012) Criticality in Large-Scale Brain fMRI Dynamics Unveiled by a Novel Point Process Analysis. Front Physiol 3 Available at: https://www.frontiersin.org/journals/physiology/articles/10.3389/fphys.2012.00015/full [Accessed July 22, 2026].

58. Van Veen BD, van Drongelen W, Yuchtman M, Suzuki A (1997) Localization of brain electrical activity via linearly constrained minimum variance spatial filtering. IEEE Trans Biomed Eng 44:867–880.

59. Wang PN, Lirng JF, Lin KN, Chang FC, Liu HC (2006) Prediction of Alzheimer’s disease in mild cognitive impairment: a prospective study in Taiwan. Neurobiol Aging 27:1797–1806.

60. Yavuz BB, Ariogul S, Cankurtaran M, Oguz KK, Halil M, Dagli N, Cankurtaran ES (2007) Hippocampal atrophy correlates with the severity of cognitive decline. Int Psychogeriatr 19:767–777.

61. Zalesky A, Fornito A, Cocchi L, Gollo LL, Breakspear M (2014) Time-resolved resting-state brain networks. Proc Natl Acad Sci 111:10341–10346.

