## Supplemental Material for "Temporal pole initiation of neuronal avalanches is associated with episodic Memory performance in Mild Cognitive Impairment"

### Supplementary Material

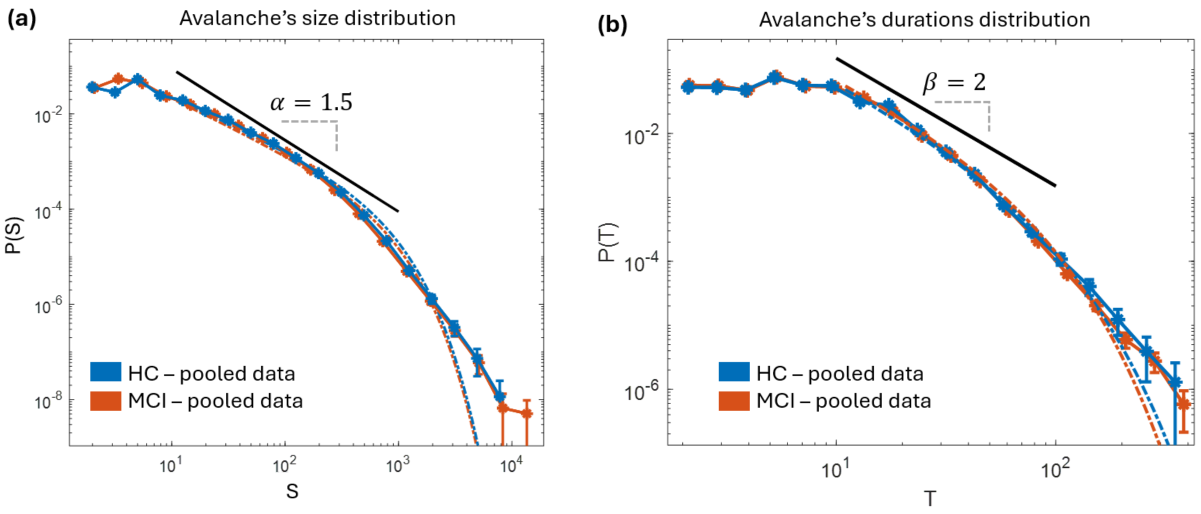


**Figure S1**. **(a)** Distribution of avalanche sizes pooled across all healthy controls (HC, blue) and patients with mild cognitive impairment (MCI, orange), displayed on logarithmic axes. Both distributions exhibit an approximately scale-free regime over an intermediate range of avalanche sizes (black line, slope corresponding to α=1.5 shown for visual guidance). The power-law exponent was estimated independently for the HC and MCI groups using (Alstott et al., 2014). When the fitting was restricted to the scaling range identified by the black line shown in the figure (S∈[10,1000]), the estimated exponents were α-HC≈1.43 and α-MCI≈1.45.
